# Prevalence of substance use and associated factors among patients with mental illness at Muhimbili National Hospital, Dar es Salaam, Tanzania

**DOI:** 10.1101/2023.01.11.23284422

**Authors:** Kilaye Karino, Joel S. Ambikile, Masunga K. Iseselo

## Abstract

The burden of substance use among patients with mental illness is prevalent in developing countries including Tanzania with negative consequences on treatment outcomes. However, the current prevalence of substance use and its associated factors in this population remains unclear in Tanzania. This study aimed at determining the prevalence of substance use and associated factors among patients with mental illness at Muhimbili National Hospital, Dar es Salaam Tanzania.

We conducted a descriptive cross-sectional study between March and May 2022. Data were collected during a face-to-face interview with patients attending the outpatient clinic at the Psychiatric Unit at MNH. We used a structured social demographic questionnaire and WHO ASSIST V3.0 tool was used to collect the data. Bivariate and multivariate regression analyses were performed using SPSS version 26 to determine the association between patient characteristics and substance use. A p-value of less than 0.05 was considered statistically significant. A total of 364 patients were enrolled. Among these, 215 (59.1%) were males and the mean (SD) age was 35.57 (±9.01) years. We found the prevalence of substance use to be 32.7%, and the most commonly used substance was alcohol (21.7%), followed by tobacco (19.8%) and cannabis (12.7%). Being a male [Adjusted Odds Ratio (AOR): 2.133; 95% Confidence Interval (CI): 1.258-3.619; P=0.005], younger age [AOR:3.301; 95% CI: 1.152-9.453; P=0.026], positive family history of mental illness [AOR:2.423; 95% CI: 1.448-4.056; P=0.001], and having a family history of substance use [AOR:3.721; 95% CI: 2.215-6.252; P=0.001], were significantly associated with substances use.

In conclusion, substance use among patients with mental illness is prevalent. Establishing a routine screening program at the psychiatric clinics is essential in identifying substance use among these risk groups and providing appropriate treatment will improve treatment outcomes. Future research should explore help-seeking behaviors and the accessibility of substance use treatment in patients with mental illness.

## Introduction

Substance use among patients with mental illness is a major challenge for mental health service providers in Tanzania and across different parts of the world [1]. Globally, half of the patients with mental illness have used substances during their lifetime, contributing to 37.6% of disease-burden-related disabilities [2]. Individuals diagnosed to have mental illness are more likely to use substances and those who use substances are more likely to experience mental illnesses [3]. The reasons for substance use among patients with mental illness are reported to be self-medication as a means of coping with symptoms, neurotransmitter changes in the brain, homelessness, and unemployment [4].

Substance use among patients with mental illness increases the risk of mortality and morbidity compared to the general population [4]. This is due to the exacerbation of mental illness with profound functional impairment. Substance use is most commonly reported in patients with anxiety disorders, personality disorders, mood disorders, schizophrenia, and post-traumatic stress disorder [5]. The most frequently used substances among patients with mental illness include tobacco, alcohol, cannabis, and cocaine [6].

In Africa, the prevalence of substance use among patients with mental illness ranges from 21.3% to 69.2% [7,8]. Moreover, substance use among patients with mental illness contributes to the high prevalence of mental illnesses and poor treatment outcomes with poor prognoses for those affected [9]. Contrary to patients who do not use substances, patients with mental illness who use substances have higher psychopathological severity with profound functional impairment, poor treatment compliance, and increased relapse, readmission, and suicide rates [10,11]. This is because substance use interferes with psychotropic medication’s beneficial effects [12].

Patients with mental illness using substances are more likely to be unemployed, homeless, and violent and have unstable family and interpersonal relationships with a diminished contribution to economic activities [13, 14]. They tend to experience victimization, social exclusion, legal problems, and a high prevalence of blood-borne infections such as HIV/AIDs or hepatitis due to their risk-taking behaviors [15]. Diagnosing substance use in patients with mental illness is difficult due to the overlap and fluctuation of symptoms, making it difficult for these patients to receive appropriate treatment [16].

In addition, acute or chronic use of substances mimics mental illness symptoms making it difficult to distinguish between symptoms due to substance use disorder or withdrawal from substance use and mental illness symptoms [17]. However, screening tools vary across countries.

Although, In Tanzania, Hauli et al in 2011 in Mwanza reported the most commonly used substances and associated factors are alcohol, tobacco, and cannabis [18]. Simon and colleagues in Dares Salaam reported moderate to high alcohol users worsen medication attitudes, adherence, and psychotic symptoms in chronic psychotic disorders [19]. However, these studies did not report the prevalence of substance use but rather focused on individual substances.

Therefore, the current study aimed at determining the prevalence of substance use and its associated factors among patients with mental illness at Muhimbili National Hospital. The findings from this study give insight into the current trend of substance use among patients with mental illness, call for routine screening of substance use among patients with mental illness, and early appropriate interventions.

## Materials and Methods

### Study design

This was a descriptive cross-sectional study that employed a quantitative approach. Cross-sectional design measures exposure and outcome in the study participants at the same time [20]. A cross-sectional design was suitable as it is relatively faster to conduct, less time-consuming, inexpensive and assesses the prevalence and associated factors among patients with mental illness unlike other study designs [21]. In addition, participants are selected based on inclusion and exclusion criteria set for the study.

### Study area and setting

The study was conducted at the adult outpatient psychiatry clinic at Muhimbili National Hospital (MNH) in Dar es Salaam, Tanzania. MNH is a national referral hospital receiving patients with mental disorders from within Dar es Salaam and from nearby regions. Psychiatry and Mental Health is one of the departments in the directorate of medical services at MNH. It provides health care for patients with mental health problems suffering from psychological, addiction, and psychiatric disorders either as inpatients or outpatients. At the time of the study, about 40-70 patients were seen in adult outpatient clinics a day. Competent Mental Health Specialists and Mental Health Nurses attend to patients at the clinic. MNH was chosen to be the convenient study site due to the availability of study participants, the convenience of time, and available resources for the timely completion of the study.

### Study population

The study population was patients with mental illness aged 18 years and older attending the adult psychiatry outpatient clinic during the study period. The study included both male and female patients who are stable.

### Inclusion and Exclusion criteria

Patients aged 18 years and above, attending the psychiatry outpatient clinic, who were diagnosed to have psychiatric disorders, and who gave consent to participate were included in the study. However, participants who had debilitating and severe mental illness or physical illnesses were excluded from this study.

### Sample size and sampling procedure

Simple random sampling was used to select the study participants. The sampling frame was established by identifying all files of patients who attended a follow-up clinic. The interviewer assigned them sequential numbers and entered them into an excel sheet. From this list, we drew a random sample using a computer-generated random numbers method. A sample of 364 participants was obtained, their clinic dates were checked, and they were approached, explained the aim of the study, the importance of their participation, risk, and benefits of the study, and participated willingly based on their respective clinic days after signing a written informed consent.

### Variables Assessed

In this study substance use among patients with mental illness was the main outcome variable. The predictor variables were the age of participants, sex, marital status, living arrangement, place of residence, employment/occupation, level of education, family history of mental illness, personal and family history of substance use, age at onset of substance use, type of substance(s) used and primary diagnosis.

### Data collection method

#### Tools and measurements

The data collection method was a face-to-face interviewer-administered questionnaire.

The interviewer administered a questionnaire was used to collect information from the respondents. We used a Kiswahili version translated from the original English version. The questionnaire consisted of socio-demographic information such as age, gender, marital status, place of residence, employment/occupation, level of education, primary diagnosis, family history of mental illness, and other information related to substance use such as personal and family history of substance use, age at onset of substance use, and type of substance(s) used.

Also, the validated Swahili-translated standardized World Health Organization (WHO) - Alcohol, Smoking, and Substance Involvement Screening Test (ASSIST) V3.0 was used to collect information on substance use. This tool has been validated for substance use screening globally (22). The tool was used to screen for substance use among patients with mental illness. It assesses a lifetime and recent (past 3 months) substance use to establish several substances and degrees of substance use and dependence and is currently used to screen for substance use at the methadone clinic at MNH with a Cronbach’s alpha of ≥ 0.80 for each of the specific substance scales and total substance participation (23). The tool consists of eight questions. The first question asked whether a respondent had ever used a given substance using a binary (yes/no) response. Responses of ‘no’ terminated further questioning about that particular substance. Questions 2-7 assessed the frequency, craving, use-related problems, others’ concerns, and control over use. The questions were repeated for any substance a respondent indicated as having used previously in question 1. Substance use was scored using the two categories (Yes or No) to obtain the prevalence of substance use among patients with mental illness.

#### Data collection procedure

The first author administered the questionnaires with the help of two research assistants that were recruited from the department of Psychiatry and Mental Health. This was because they were competent and experienced in working with patients with mental illness for many years and had prior experience in research activities. To ensure compliance with the current study, the research assistants were trained on study objectives, data collection tools and how to administer them, the data collection process, and ethical procedures for the study. On the day of the interviews, ten to twelve participants were interviewed and each interview lasted 12 to 20 minutes. The interviews were conducted only during the clinic days. The files of interviewed participants were labelled ‘interviewed’ on a particular date to avoid repeating interviewing the same participant. Moreover, every patient and relative was asked before the interview if they had already been interviewed. The filled questionnaire was cross-checked for completeness before the participant left the interview room. The procedure continued daily until the desired sample was attained.

### Data analysis

Data were entered, cleaned, and processed using SPSS version 26. Descriptive statistics were summarized using mean and standard deviation for continuous variables while percentages and frequency were used for categorical variables. Bivariate analysis was performed to determine the association between socio-demographic characteristics and substance use. Odds ratios (OR) and 95% confidence intervals (CI) were used as measures of association. Variables such as age, sex, marital status, education level, living arrangement, family history of mental illness, and substance use were entered in a multivariate logistic model regression model to determine true association. A p-value <0.05 was considered statistically significant.

### Ethical consideration

Ethical approval was obtained from the Research Ethics Committee of Muhimbili University of Health and Allied Sciences (MUHAS) with reference number MUHAS-REC-03-2022-1041. Permission to conduct the study was obtained from the Teaching, Research, and Consultancy Unit of Muhimbili National Hospital and the head of the Department of Psychiatry and Mental Health. Participants with their accompanying relatives have fully explained the aim, nature, benefit, and risk of the study. Moreover, the right to withdraw from the study was clearly described. They were also explained that participation was voluntary. Participants were ensured confidentiality by using the anonymity of the information in all documents related to participant Study participants signed informed consent before data collection

## Results

### Socio-demographic characteristics and mental illness of the study participants

The total number of study participants was 364. The majority, 215 (59.1%) of participants were males. The mean age (STD) was 35.57 (9.01), the majority (171; 47%) were in the age group 25-34 years, and more than half (55.5%) were single. Of the total participants, 143 (39.3%) attended primary education while 204 (56.3%) were unemployed. Only 312 (85.7%) of the participants were living with their family members with 125 (34.3%) and 140 (38.5%) reporting a family history of mental illness and substance use respectively. The majority of 237 (65.1%) participants were diagnosed to have schizophrenia (**Table 1)**.

**Table 1:** Characteristics of patients with mental illness at MNH (n=364)

| Variables | Frequency (n) | Percentage (%) |
| --- | --- | --- |
| **Age (years)** |  |  |
| 18-24 | 31 | 8.5 |
| 25-34 | 171 | 47 |
| 35-44 | 106 | 29.1 |
| ≥45 | 56 | 15.4 |
| **Sex** |  |  |
| Male | 215 | 59.1 |
| Female | 149 | 40.9 |
| **Marital status** |  |  |
| Single | 259 | 71.2 |
| Married | 41 | 11.3 |
| Separated | 47 | 12.9 |
| Widow | 17 | 4.7 |
| **Education level** |  |  |
| No formal education | 16 | 4.4 |
| Primary school | 143 | 39.3 |
| Secondary school | 155 | 42.6 |
| Tertiary | 50 | 13.7 |
| **Occupation** |  |  |
| Employed | 17 | 4.7 |
| Self-employed | 142 | 39.0 |
| Unemployed | 205 | 56.3 |
| **Living arrangement** |  |  |
| Living alone | 22 | 6.0 |
| Living with friends | 21 | 5.8 |
| Living with family | 296 | 81.3 |
| Living with a partner/spouse | 25 | 6.9 |
| **Reported a family history of mental illness** |  |  |
| Yes | 125 | 34.3 |
| No | 238 | 65.4 |
| **Reported a family history of substance use** |  |  |
| Yes | 140 | 38.5 |
| No | 224 | 61.5 |
| **Mental illness** |  |  |
| Bipolar Mania | 30 | 8.2 |
| Cannabis induced psychosis | 12 | 3.2 |
| Major depressive disorder | 61 | 16.8 |
| Posttraumatic stress disorder | 10 | 2.7 |
| Schizoaffective disorder | 14 | 3.8 |
| Schizophrenia | 237 | 65.1 |

### Prevalence and types of substances used among patients with mental illness

The overall prevalence of substance use was 32.7% **(Figure 1)**

**Figure 1:**
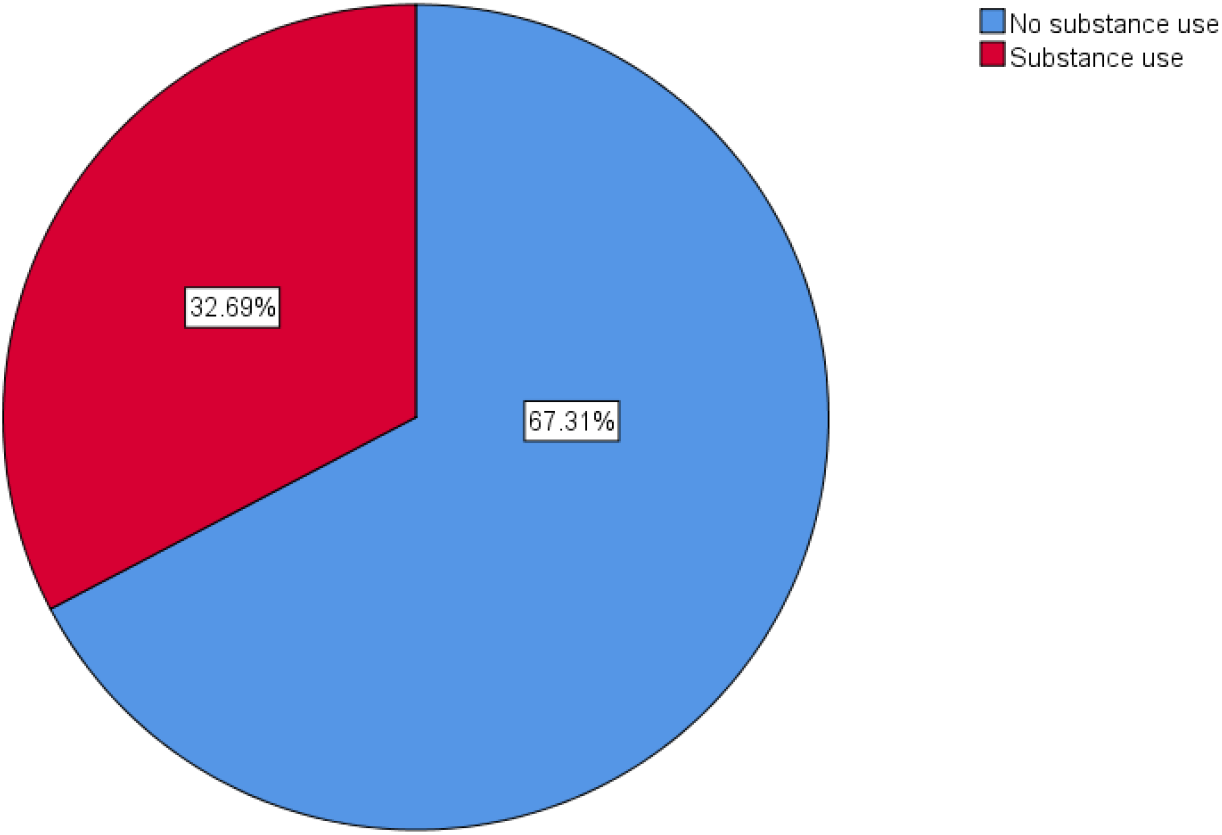
Prevalence of substance use among patients with mental illness at MNH (n=364)

The most common types of substances used among 364 participants were alcohol (21.7%), followed by tobacco (19.8%), and cannabis (12.8%). (**Figure 2)**

**Figure 2:**
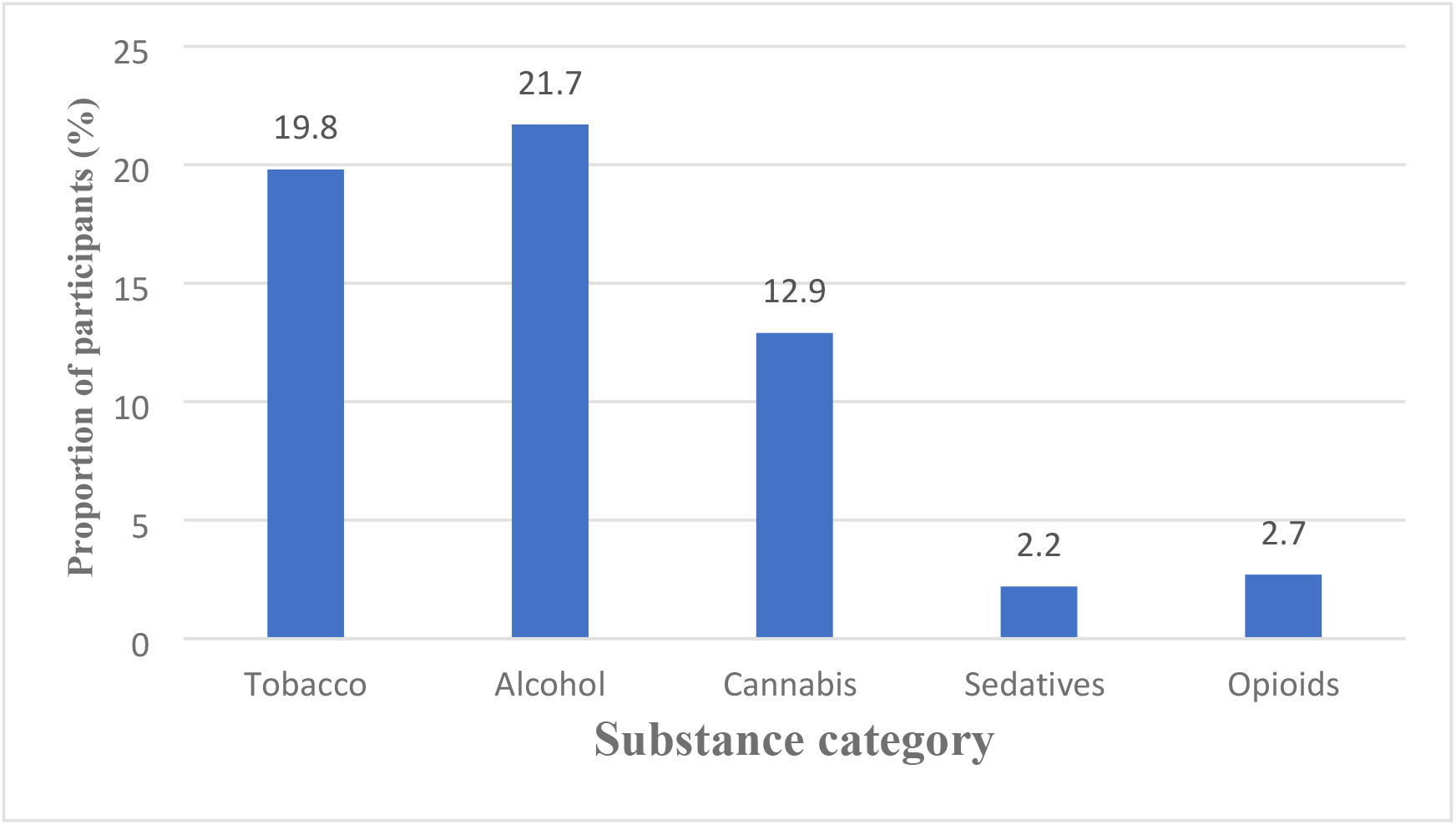
Proportion of substance used categories among patients with mental illness attending the psychiatry OPD clinic at MNH (n=364)

### Factors associated with substance use among patients with mental illness

Participants aged 18-24 had three times higher odds of using substances (COR:3.036; 95% CI: 1.216-7.576; P=0.017) compared to those aged more than 45 years. Being a male (COR:1.866; 95% CI: 1.175-2.964; P=0.005) increased the odds of using substances compared to being a female. Moreover, participants who reported a positive family history of mental illness had 3 times the odds of using substances (COR:2.622; 95% CI: 1.658-4.145; P=0.01 compared to those with no family history of substance use. The participants having a family history of substance (s) had four times higher odds of using substances (COR:3.773; 95% CI: 2.382-5.977; P=0.01) compared to those without a family history of using a substance (s).

However, after adjusting for the confounders, participants of younger age (18-24) had higher odds of using a substance(s) (AOR:3.301; 95% CI: 1.152-9.453; P=0.026) than the older groups. Males had two times higher odds of using a substance(s) (AOR:2.133; 95% CI: 1.258-3.619; P=0.005) compared to females. Participants with a positive family history of mental illness were two times more likely to use a substance(s) (AOR:2.423; 95% CI: 1.448-4.056; P=0.001) compared to those without a family history of substance use. Moreover, those who reported a family history of substance(s) use were four times more likely to use the substance(s) (AOR:3.721; 95% CI: 2.215-6.252; P=0.001) compared to their counterparts **(Table 2)**.

**Table 2:**
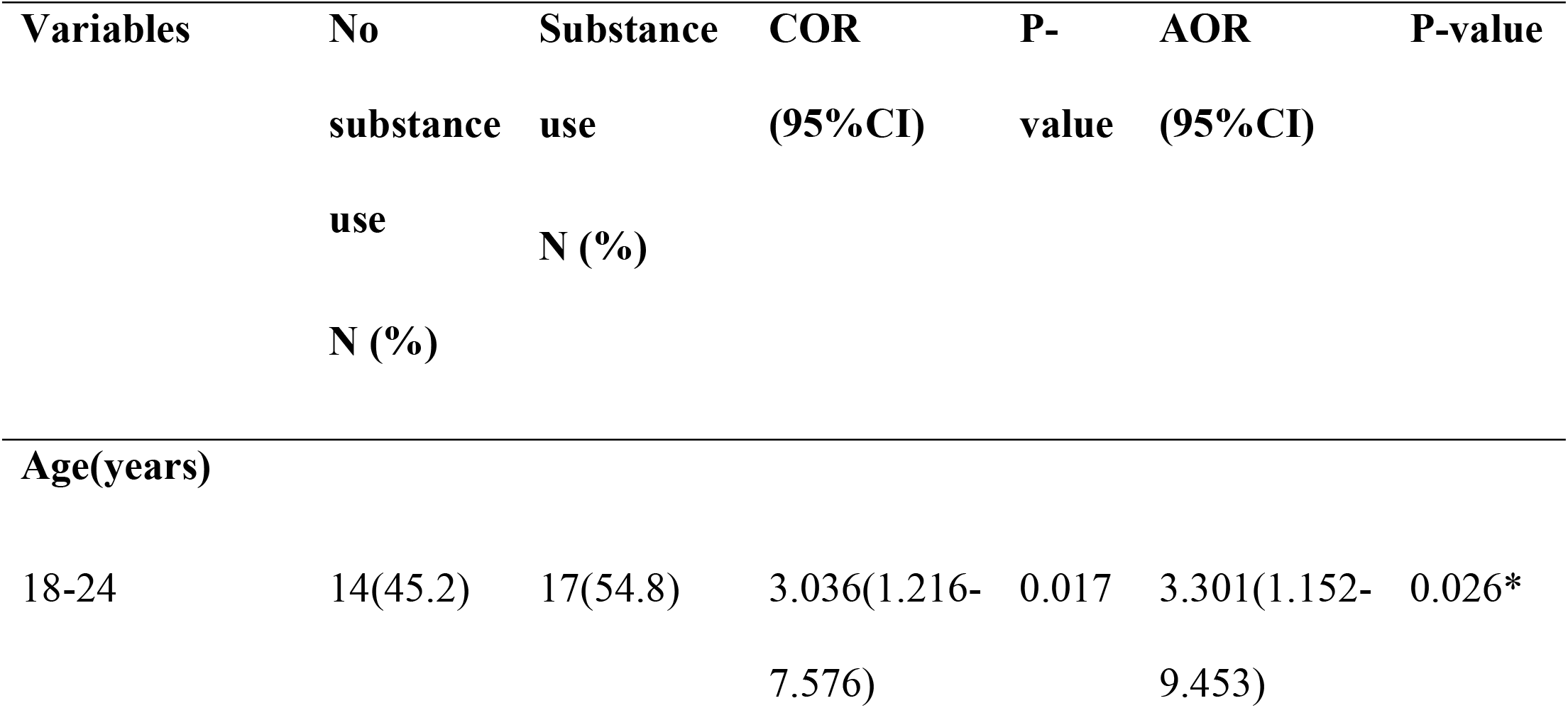
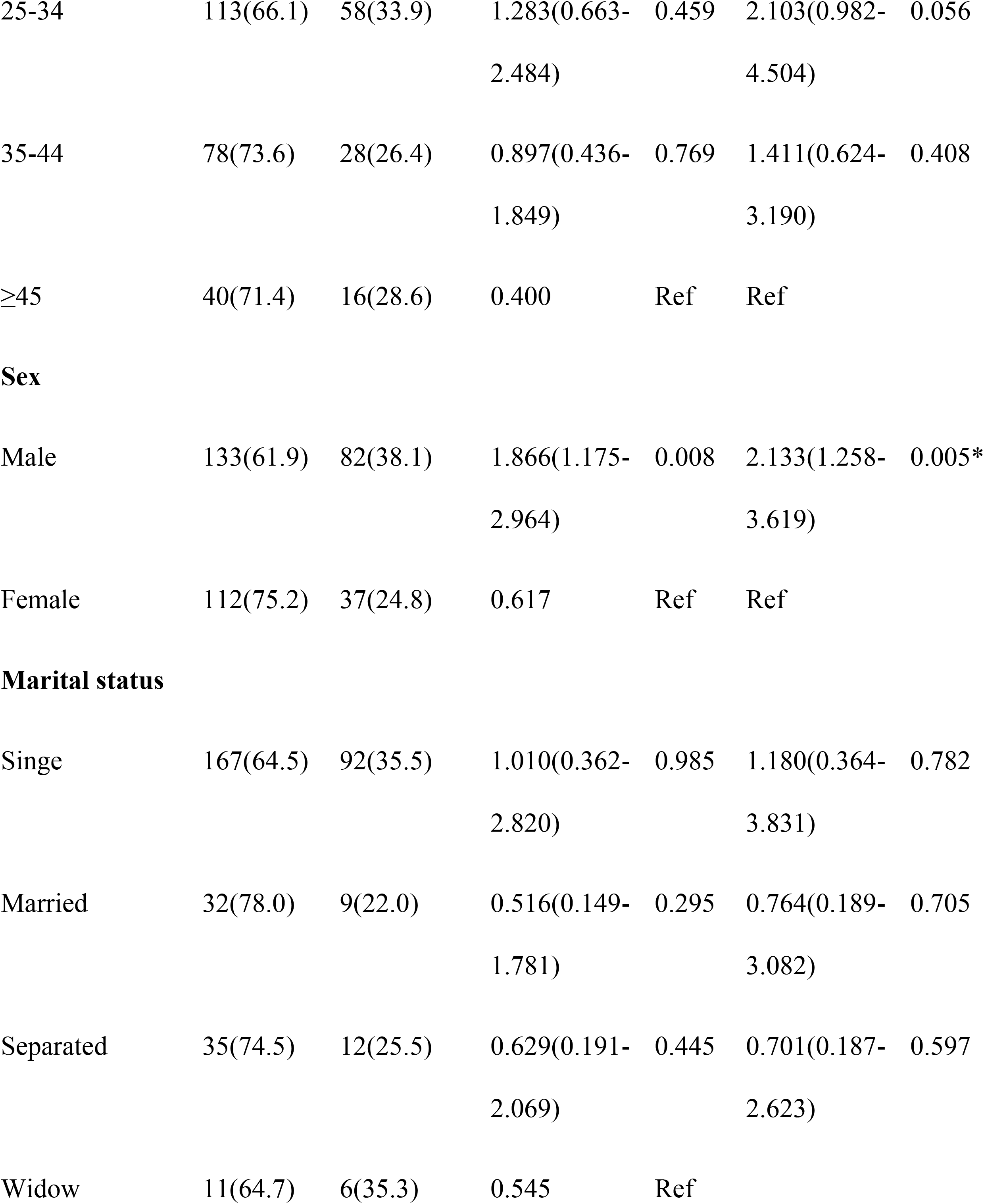
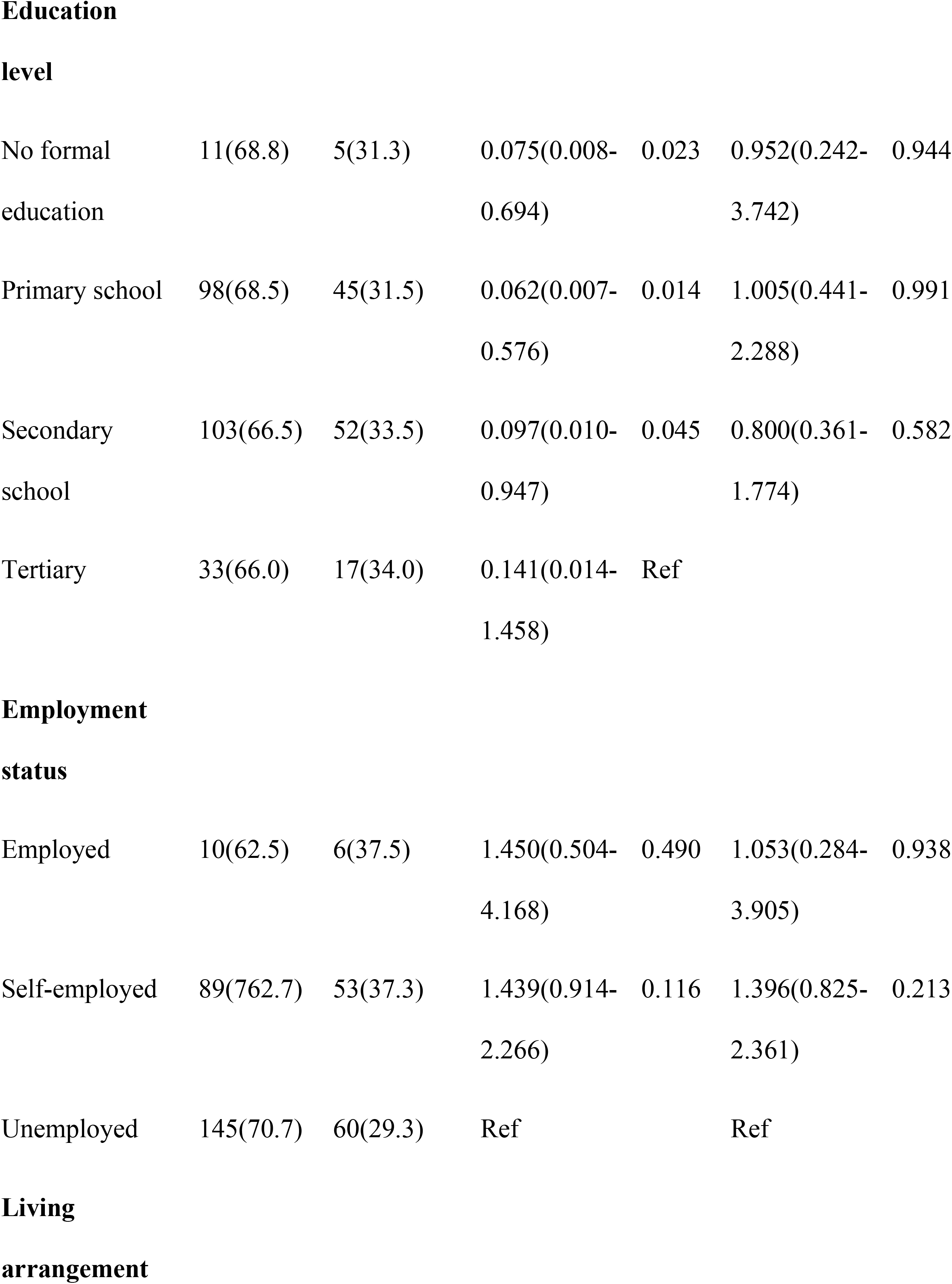
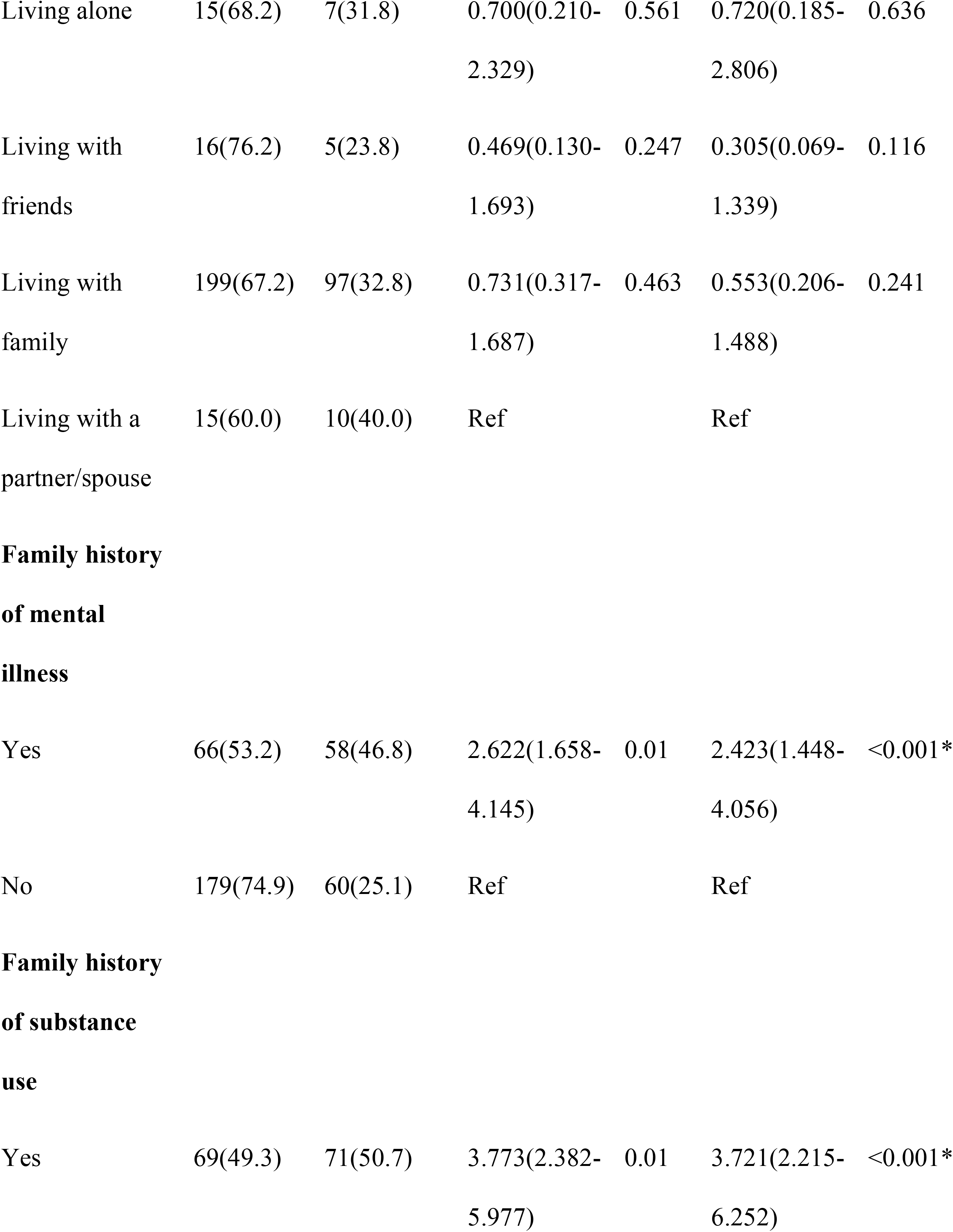
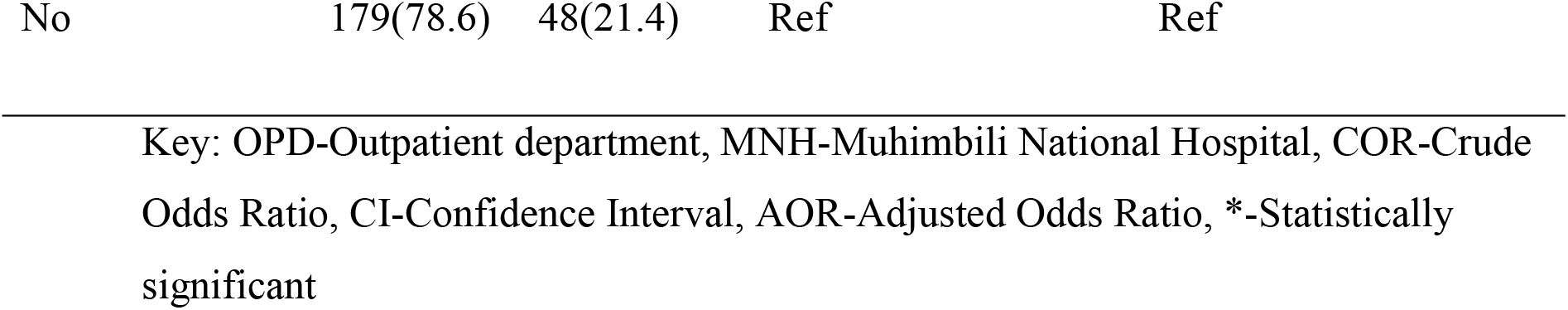
Association between participants’ characteristics and substance use among patients with mental illness attending the psychiatric OPD clinic at MNH (n=364)

## Discussion

This study aimed at determining the prevalence of substance use and associated factors among patients with mental illness who attended psychiatry outpatient clinics at MNH. The prevalence of substance use was found to be higher among patients with mental illness. The most commonly used substances include alcohol, tobacco, and cannabis. Being a male, younger age, having a family history of mental illness, and history of substance use in the family were significantly associated with substance use in patients with mental illness.

### Prevalence of substance use among patients with mental illness

Our results indicated a high prevalence (32.7%) of substance use among patients with mental illness. This prevalence shows that the risk of substance use is high among mentally ill patients. However, substance use in patients with mental illness remains underdiagnosed contributing to an increased risk of relapses due to poor medication adherence. This indicates a need for routine screening for early detection of substance use in these patients and for establishing appropriate measures.

The prevalence of substance use in our study is low compared to the study reported in a similar cross-sectional study in Namibia [24] and a tertiary hospital in Iran [3] and India [7]. This variation in prevalence might be due to sampling size differences and cultural norms. However, the prevalence in our findings was higher than that observed among patients with mental illness in South Africa, the US, Malaysia, and Finland [9,25,26]. This may be explained by variations in access to mental health services and poor health-seeking behavior, especially in Africa, which results in using the substance of abuse as a treatment option or coping with the illness. Moreover, this may as well be attributed to differences in the perception of illness and treatment where some don’t believe in antipsychotics and opt for substances as the best treatment option [27].

In the current study, patients with mental illness had a significantly higher risk of substance use than the general population because common risk factors can contribute to both mental illness and substance use. This shows a need for special attention to this particular risk group to identify substance use early and treat it appropriately. However, our findings are similar to a study conducted in Norway [28]. Screening for substance use and treatment of both mental illness and substance use problems improve medication adherence and reduction relapses as a consequence of substance use among this population.

### Commonly used Substances among patients with mental illness

Tobacco, alcohol, and cannabis were the common substances used by the participants in our study. This could be due to the easy availability and accessibility of these substances. Tobacco is produced in Tanzania under Tanzania Cigarette Company, a subsidiary of Japan Tobacco International which dominates the tobacco market by 96.1% along with British American Tobacco by 1.3%, and Philip Moris International by 1.2% [29]. The tobacco products regulation act 2003 regulates the manufacturing, labelling, distribution, sale, and advertising of tobacco products and smoking in public spaces [30].

Cannabis is illegal in Tanzania, unlike alcohol which is easily accessible in local bars. It’s not easy to know if cannabis and other illegal drugs exist unless someone is in the loop of business which is done in secrecy [31]. These findings were relatively similar to those reported in a cross-sectional study in lake zone Tanzania whereby [18] and other countries such as Sweden [36], Norway [17,32,33], and Nigeria [7]. Similarities in the type of substance (s) commonly used among patients with mental illness between the current study and other studies can be explained by the availability of the particular substances in those countries and the patient’s characteristics. Patients with mental illness experience stressors associated with illness, and neurochemical changes in the brain which might increase the need for substances such as alcohol, cannabis, tobacco, and others [34]. Using these particular substances among patients with mental illness can be explained by the need for self-medication to deal with negative symptoms such as social withdrawal, apathy, dysphoria, and sleep problems as well as to decrease the discomfort from the side effects of antipsychotic medication [35].

Conversely, the current study findings differ from a systematic review that reported that benzodiazepines and opioids were the most frequently used substance among patients apart from cannabis [34], and in India whereby the most common substance used were tobacco apart from alcohol and cannabis [37]. Moreover, different types of substances were reported in the US such as cocaine, hallucinogens, opioid, methamphetamine, and tobacco [38], stimulants in Denmark [39] and cough syrup, and codeine in Nigeria [4]. Again, this variation in the type of substances used can be explained by the availability of the substance in a certain geographical area and the social demographic characteristics of the study participants. Substance use among patients with mental illness complicates treatment outcomes and results in an increased risk of relapse. Education on substance use-related problems and their impact on treatment outcomes are highly essential [22].

In our study, substance use was common among patients with schizophrenia contrary to that in Denmark where substances, particularly stimulants and cannabis, were frequently used among patients with bipolar disorders [37]. The variation in types of substance use among patients with mental illness between our study and other studies might be due to willingness to report the type of substance being used, cultural norms, and legal frameworks of the particular locality.

### Factors associated with substance use among patients with mental illness

The current study found that being a male, of younger age, having a positive family history of mental illness and family history of substance use were significantly associated with substance use among patients with mental illness. Routine screening among these at-risk groups would result in early diagnosis, treatment, and improved treatments outcome. Our findings are consistent with a report by Karpov et al. in Finland in 2017 [26]. However, our findings are contrary to the results of a study in Malaysia and Nigeria which demonstrated that younger individuals particularly men, brought by police and those diagnosed to have schizophrenia, being married with at least primary education, and being unemployed were significantly associated with an increased risk of substance use [7, 25].

The current study reported that males have 2 times higher odds of substance use than females. Similar findings have been previously reported in the same country (Mwanza region), the US, Norway, Denmark, and Nigeria [9, 17, 18, 41, 42] showing that being a male is a significant predictor of substance use among patients with mental illness. This is explained by the possibility that sex significantly influences a particular type of mental disorder an individual develops, the differences in socio-economic determinants of mental health, social status, susceptibility, and exposure to life stressors and coping skills. Males are more likely to use substances due to early life peer pressure, curiosity, and exploring an environment where substances are available [43].

This study has shown that younger age increases the odds of substance use among patients with mental illness. Similar findings have also been reported in Nepal, Nigeria and South Africa [7, 44, 45]. Substance use among younger individuals might be due to peer pressure, exposure to stressors with underdeveloped coping skills, the curiosity of wanting to experience the effects of a particular substance, early use, and lack of family guidance at an earlier age [34]. On the contrary, other studies have shown factors such as marital status, occupation, duration of illness, owning a business, and being unemployed to be significantly associated with higher odds of substance use [45, 43]. This contradiction could be due to differences in social-demographic characteristics of study participants such as age.

Family history of mental illness and family history of substance use were significantly associated with substance use in the current study. These findings are in line with those previously reported by Hauli et al and Khokhar et al. in the same country and in the US, respectively [46, 18]. This may be explained by the fact that common risk factors contribute to substance use and mental illness although the exact mechanism is unknown. People who use substances are more likely to experience mental illness and vice versa [27]. In our study, participants with a positive family history of mental illness were twice more likely to use substances while those with a family history of substance use were 4 times more likely to use substances compared to their counterparts. This indicates a true association between positive family history of mental illness and substance use among patients with mental illness.

Therefore, substance use among patients with mental illness younger age, being male, and having a family history of mental illness and substance was seen as common predictors of substance use among patients with mental illness.

### Study limitations and mitigation

This was a cross-sectional study, and can, therefore, not establish a cause-effect relationship as it assessed substance use and its associated factors at the same time. Social desirability bias could also happen since the study was based on self-reports of substance use. Participants might have reported what they thought the interviewer wanted to hear, or what was perceived as socially acceptable, which would have underestimated or overestimated the prevalence obtained in this study. In mitigating these limitations, accompanying relatives confirmed whether the information provided by the patients was correct. In addition, participants were ensured confidentiality to promote free expression and comfort.

## Conclusions

This study unveils the prevalence and associated factors of substance use among patients with mental illness in Tanzania. Our study demonstrated that patients with mental illness who use the substance (s) remain unrecognized thus complicating treatment outcomes. The study underscores the need for substance use awareness programs, routine screening, early identification, and appropriate treatment of substance use in patients with mental illness. Mental health care providers should establish education awareness sessions or programs in the clinics and use local media to disseminate information on the effect of substance use among patients with mental illness. Future studies should explore help-seeking behaviors and the accessibility of substance use treatment in patients with mental illness.

## Data Availability

All relevant data are within the submitted manuscript

## Acknowledgment

We would like to thank all the participants for allowing us to conduct the interview. We acknowledge the research assistant involved in the data collection process, Ms Mary Njau and Mr Chundi George. Finally, we thank the Muhimbili National Hospital administration for permission to conduct this study.

## Authors contribution

**Conceptualization:** Kilaye Karino, Masunga K. Iseselo, Joel S. Ambikile

**Data curation:** Kilaye Karino, Masunga K. Iseselo, Joel S. Ambikile

**Formal analysis:** Kilaye Karino

**Investigation:** Kilaye Karino

**Methodology:** Kilaye Karino, Masunga K. Iseselo

**Supervision:** Joel S. Ambikile, Masunga K. Iseselo

**Validation:** Joel S. Ambikile

**Visualization:** Kilaye Karino, Joel S. Ambikile

**Writing-original draft:** Kilaye Karino

**Writing-review & editing:** Masunga K. Iseselo, Joel S. Ambikile

